# Drawing transmission graphs for COVID-19 in the perspective of network science

**DOI:** 10.1101/2020.08.11.20172908

**Authors:** N. Gürsakal, B. Batmaz, G. Aktuna

## Abstract

When we consider a probability distribution about how many COVID-19 infected people will transmit the disease, two points become important. First, there should be super-spreaders in these distributions/networks and secondly, the Pareto principle should be valid in these distributions/networks. When we accept that these two points are valid, the distribution of transmission becomes a discrete Pareto distribution, which is a kind of power law. Having such a transmission distribution, then we can simulate COVID-19 networks and find super-spreaders using the centricity measurements in these networks. In this research, in the first we transformed a transmission distribution of statistics and epidemiology into a transmission network of network science and secondly we try to determine who the super-spreaders are by using this network and eigenvalue centrality measure. We underline that determination of transmission probability distribution is a very important point in the analysis of the epidemic and determining the precautions to be taken.

## INTRODUCTION

The first half of 2020 passed with the whole world dealing with the COVID-19 outbreak. First, many countries implemented lockdown, and then reopening came to the agenda. However, at the time of writing this article, there was an important increase in the number of infections all over the world and we would probably spend the second half of the year dealing with the COVID-19 issue and lockdowns again. The fact that COVID-19 is a relatively new virus also challenges scientists and scientific analysis have to navigate the uncharted territories [1].

This article attempts to establish a link between the fields of statistics, network science and epidemiology using an interdisciplinary approach. From a micro point of view, this connection, which was tried to be established, was made by converting transmission distribution of statistics and epidemiology into a transmission network of network science. From a micro point of view, the study also tries to contribute to efforts to stop the epidemic by identifying who the super spreaders are, and then by researching and identifying their various characteristics.

In analyzing COVID-19 outbreak, most of the times, instead of focusing on a transmission probability distribution; R_0_ value as an average or median have been used and super-spreaders are not taking into account. But the extreme values make a long tail for this distribution and rare infection events determine the shape of this distribution.

It has been seen that different R_0_ numbers were obtained in the literature for COVID-19 and some authors try to emphasize that 80% of cases are infected by a group of 20%. But most of the times, the fact that 80% of the infection is carried out by a 20% group is often not considered and most of the analysis begin with a R_0_ reproduction number.

Nearly all of the COVID-19 studies begin with the determination of reproduction number R_0_. For example, if a disease has an R_0_ of 15, a person who has the disease will transmit it to an average of 15 other people. This coefficient is important because also we use this number to see the severity of outbreak and this number is also used as an epidemic threshold parameter.

Different methods are being used to determine this number. First of all, this number can be calculated as:

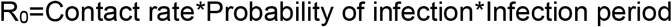

Secondly, this coefficient can be calculated using “attack rate” and attack rate is the percentage of the population eventually infected.

Thirdly we can estimate R_0_ using formula:

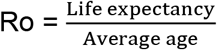

And fourthly we can estimate R_0_ using exponential growth rate.

“Since the R_0_ has a key role in measuring the transmission of diseases and is crucial in preventing epidemics, thus it is important to know which methods and formulas to apply to estimate R_0_ and have better performance” [2]. But we know that different methods give different results [3] and most of the times in scientific articles which method has been used is not mentioned. Besides, sometimes R_0_ value is given as a median; for example, it is expressed as, “We estimated that the median of estimated R_0_ is 5.7 (95% CI of 3.8-8.9)” [4] and this may lead us to some confusion too.

“The emerging picture for epidemic spreading in complex networks emphasizes the role of topology in epidemic modeling” [5]. Disease transmission networks have the motifs of transmission stories. One of the most important ways to avoid contamination is to have information about how this transmission happens.

The main purpose of this study is to develop a simple method that will make it easier for us to look at the COVID-19 issue from the network science window. And focusing on the interplay between network theory, statistics and epidemiology [6] In this simple method, first we determine a transmission probability distribution and secondly simulating this probability distribution we can draw a transmission graph and try to understand the process contamination using this graph. We should underline that determination of transmission probability distribution is a very important point in the analysis of the epidemic and determining the precautions to be taken. Having such a graph, also we may compute many network measures and use this measures in our decision process.

Our study, which started with an introduction, includes a literature review on the R_0_ values calculated for COVID-19. The following section examines the phenomenon of super-spreaders. Then, the issues of generating the values obtained by simulating a discrete Pareto distribution and drawing and interpreting the transmission network are included. The study is completed with a discussion section.

### R_0_ values calculated for COVID-19

COVID-19 transmission is strongly heterogeneous [7]. A high degree of individual-level variation in the transmission of COVID-19 has been expressed and consensus range *of R_0_* was found within the interval of 2-3 [8]. Results show that there was probably substantial variation in SARS-CoV-2 transmission over time after control measures were introduced. In the beginning of outbreak in Wuhan R_0_ median daily reproduction number changes from 2.35 to 1.05 in only one week. But even in the before the travel restrictions period it was found that, median daily R_0_ changes between 1.6 and 2.6 in Wuhan [9]. In the early days of outbreak in Wuhan another study estimates R at 0.24 (95% CrI: 0.01-1.38) for market-to-human transmission and 2.37 (95% CrI: 2.08-2.71) for human-to-human transmission [10]. And also in another paper it was stated that, “We identified four major clusters and estimated the reproduction number at 1.5 (95% CI: 1.4-1.6)” [11].

“We modelled the transmission process in the Republic of Korea and Italy with a stochastic model, and estimated the basic reproduction number R_0_ as 2.6 (95% CI: 2.3-2.9) or 3.2 (95% CI: 2.9-3.5) in the Republic of Korea, under the assumption that the exponential growth starting on 31 January or 5 February 2020, and 2.6 (95% CI: 2.3-2.9) or 3.3 (95% CI: 3.0-3.6) in Italy, under the assumption that the exponential growth starting on 5 February or 10 February 2020, respectively.” [12]. From these lines we learn that R_0_ is between 2.6-3.2 for Republic of Korea and 2.6-3.3 for Italy.

Transmission was modelled as a geometric random walk process, or negative binomial offspring distribution is used to calculate the probabilities: “Once we had estimated R_t_, we used a branching process with a negative binomial offspring distribution to calculate the probability an introduced case would cause a large outbreak” [9]. Also, “The relative odds of transmission among contacts of various types were estimated by use of conditional logistic regression” [13].

Another important study, in a Bayesian framework tries to use a matrix of transmission probabilities to estimate the transmission network and in this article transmission probabilities have been estimated by Monte Carlo Markov Chain procedure using Metropolis-Hastings algorithm [14].

### Super-Spreaders

We want to give three examples to show how the same virus can have different results in different environments. We should also add that there are no big differences in the dates of the events as seen from footnotes:

> “For example, the value of R on the Diamond Princess cruise ship was estimated to be 11 even though the worldwide average is 3.28. The close confines and movement of the ship’s staff facilitated COVID-19 transmission. The virus was the same, but the environment and behavior were different, altering R of the virus.”[15]
>
> “Since then, epidemiologists have tracked a number of other instances of SARS-CoV-2 super-spreading. In South Korea, around 40 people who attended a single church service were infected at the same time.”[16]
>
> “At a choir practice of 61 people in Washington state, 32 attendees contracted confirmed COVID-19 and 20 more came down with probable cases. In Chicago, before social distancing was in place, one person that attended a dinner, a funeral and then a birthday party was responsible for 15 new infections.” [16]

Looking at the outbreaks in history, it can be seen that the phenomenon of super-spreaders is not new. “We examine the distribution of fatalities from major pandemics in history (spanning about 2,500 years), and build a statistical picture of their tail properties. Using tools from Extreme Value Theory (EVT), we show for that the distribution of the victims of infectious diseases is extremely fat-tailed” [17]. Susceptible hosts within a population had not equal chances of becoming infected. Although “it is still unclear why certain individuals infect disproportionately large numbers of secondary contacts” [18]. If we have extreme transmitters, then “the practice of relying on an average R_0_ in dynamic disease models can obscure considerable individual variation in infectiousness”[19].

Heterogeneities in the transmission of infectious agents are known since the end of 1990’s [20]. We can define the phenomenon of super-spreaders in the framework of network science as follows: “The super-spreaders are the nodes in a network that can maximize their impacts on other nodes, as in the case of information spreading or virus propagation “ [21]. This definition reminds us the outlier concept of statistics, But super-spreaders are not outliers that can be discarded from analysis, “In this framework, super-spreading events (SSEs) are not exceptional events, but important realizations from the right-hand tail of a distribution” [22].

Another definition starts with establishing the relationship between heterogeneity and super-spreaders: “Heterogeneity in R_t_ between cases appears to be particularly important for SARS because of the occurrence of “super-spread events (SSEs)”—rare events where, in a particular setting, an individual may generate many more than the average number of secondary cases” [23].

One of the most important features of COVID-19 in contamination in society is the Pareto principle created by super-spreaders. Super-spreaders transmit the disease to a large number of people in an outlier-like manner, resulting in few people transmitting the disease to a large number of people, and as a result we have a transmission distribution as a power-law distribution.

Researchers are beginning to come to a consensus that coronavirus transmission more or less follows the 80/20 Pareto Principle [24] and estimated that 20% of cases were responsible for 80% of local transmission [25]. When we examine the researches, we can see that some results have slightly different results than the Pareto Principle such as, “indicated high levels of transmission heterogeneity in SARS-CoV-2 spread, with between 17-10% of infected individuals resulting in 80% of secondary infections” [26] and “Model suggested a high degree of individual-level variation in the transmission of COVID- 19…suggesting that 80% of secondary transmissions may have been caused by a small fraction of infectious individuals (~10%)” [8]

## METHODS

In the outbreak, the number of patients’ variable, in other words, the number of transmissions changes only at the discrete points of time. This means that the number of transmissions in outbreaks is discrete variables. The number of COVID 19 patients varies in many situations; how many new people are infected with the disease, how many patients have recovered and how many of them are likely to be infected or infectious again. If we were able to predict the transmission processes, there would be a deterministic approach in creating a transmission network however its impossibility is clear. Therefore, this is a stochastic process, as the number of transmission and from whom it is transmitted cannot be fully estimated or determined. Probability distributions are used in stochastic simulation models. In the simulation model, the number of transmissions, which is an discrete event, varies in the number of patients depending on time [between ‘t’ time point and ‘t + 1’ time point (s ‘t’ and ‘t + 1 may be, 7 days or 14 days period)] which is a change or increase is a discrete value and this change or increase is a discrete value. However, since there is no definite vaccination or herd immunity, person-to-person transmission remains a dynamic process.

In simulation models, the distribution is determined by collecting information and data about the subject studied. In the light of these data, the model is established by determining the probability distributions of probabilistic (stochastic) processes. In this study, as a result of literature search, the principle that COVID-19 transmission is related to Pareto distribution and power-law has been adopted and we decided the distribution to be produced in the drawing of the random transmission network, as the discrete Pareto distribution. By simulating the Pareto distribution, disease transmission data from one patient to others was randomly derived. The network drawn with the derived data is created according to the Power-low distribution. Power-low is independent of scale. The concept of independence from the scale indicates that the ratio and probability in small numbers such as 10, 40 are equal to the ratio and probability in large numbers such as 1000, 40000. In such networks, few nodes have many connections, and many nodes have few connections the “rich get richer” rule (preferential attachment,) remains valid in connection [27]. The same similar logic has been applied in the determination of super-spreaders in drawing the COVID-19 network. Thus, the study is based on the principles that some people infect more people(super- spreaders) and some infect less or not (isolate).

## RESULTS

### Simulating Discrete Pareto Distribution (Zipf Distribution)

Zipf, Pareto and power law “terms are used to describe phenomena where large events are rare, but small ones quite common. For example, there are few large earthquakes but many small ones. There are a few mega-cities, but many small towns. There are few words, such as ‘and’ and ‘the’ that occur very frequently, but many which occur rarely” [28]. Economists know that Wilfried Fritz Pareto observed that 20% of Italians held 80% of the country’s wealth in the 19^th^ century. Pareto principle also known as the 80/20 rule.

If we give an example of one of the studies on the transmission distribution as power-law, “The empirical data are highly consistent with the hypothesis that the number of reported cases are taken from a truncated power-law distribution of the form P(n) ~ n^-μ^, 1 ≤ n ≤ n_max_ “ [29]

Using the code in an R source [30], we can simulate a discrete Pareto distribution and draw its histogram as in Figure 1.

> library(degreenet)

> a=simdp(n=1000,v=2.5, maxdeg=40)

> head(a,100)

[1] 1 1 3 2 1 1 1 1 2 1 1 2 2 1 1 1 1 1 1 4 2 2 1 2 1

[26] 1 1 2 1 2 11 1 2 1 1 1 3 1 3 3 1 1 1 1 1 1 1 2 1 1

[51] 1 1 1 1 1 1 1 1 1 1 1 9 1 4 11 1 1 2 1 1 1 1 1 1 1

[76] 1 1 2 1 1 1 1 1 1 2 1 1 1 2 1 1 1 1 2 1 3 1 8 1 4

> hist(a,breaks=50)

**Figure 1.**
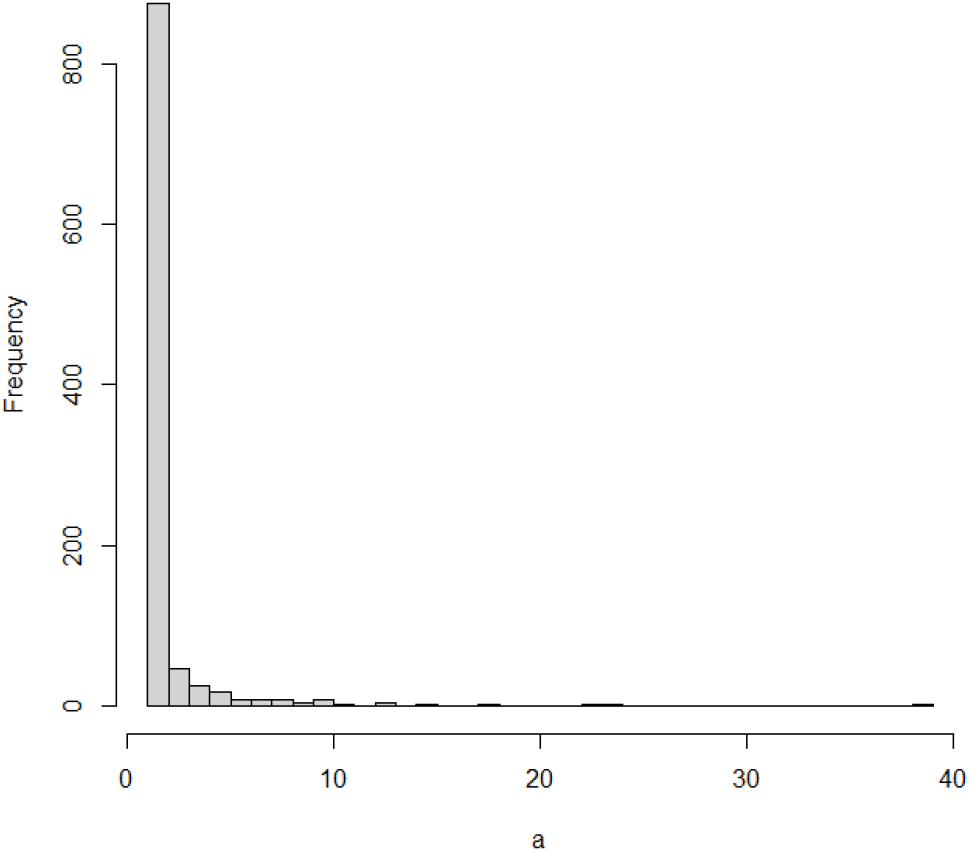
Histogram of a simulated discrete Pareto distribution

### Drawing COVID-19 Transmission Graphs

Contact networks and disease transmission networks are different from each other. However, although data on this subject is obtained through filiation studies or contact trace apps, it is not easy to convert these data into social network-like networks due to some uncertainties and bureaucratic problems. In this case, how to determine a synthetic COVID- 19 transmission network question becomes important.

The number of incoming connections in a “contact” network does not have to be one. For example, in Figure 2, people B and D were placed closer than 1.5 meters from C, but the people which transmitted disease to C, was B, as seen in Figure 2. Briefly, nodes in contact networks can have multiple incoming and outgoing connections.

> library(igraph)

> el <- cbind(c(“A”,”B”,”D”,”E”,”E”,”E”,”A”,”D”),

+ c(“B”,”C”, “A”,”F”,”G”,”H”,”E”,”C”))

> g <- graph.edgelist(el, directed=TRUE)

> plot(g)

**Figure 2.**
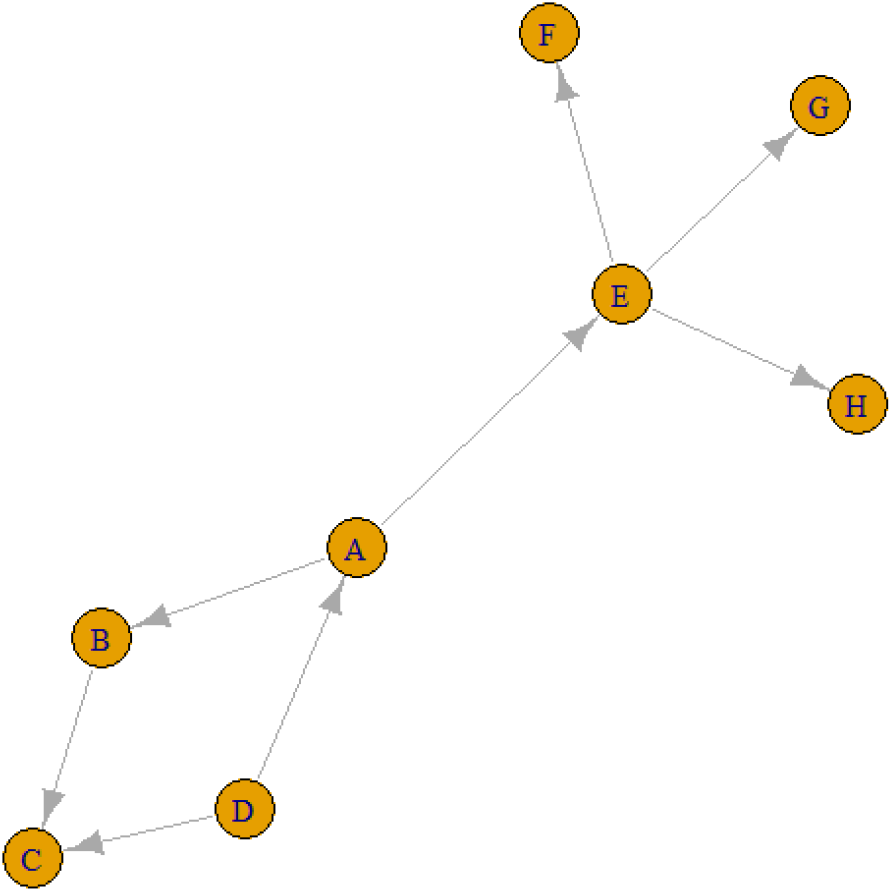
A contact graph

Eventually, “contact” networks become “contamination” networks. All these graphs, come from “contact” networks. The graph in the Figure 2 is a “transmission” graph, and as can be seen, the number of incoming connections are one for all these 5 nodes. In contrast, the number of outgoing connections can be an integer greater than one. For example, in the Figure 3, E is infected by A and F, G and H are infected by E. Similarly, A is infected by D, and B and E are infected by A. These can be referred to as “transmission” graphs.

> library(igraph)

> el <- cbind(c(“A”,”B”,”C”,”D”,”E”,”E”,”E”,”A”),

+ c(“B”,”C”, “D”, “A”,”F”,”G”,”H”,”E”))

> g <- graph.edgelist(el, directed=TRUE)

> plot(g)

**Figure 3.**
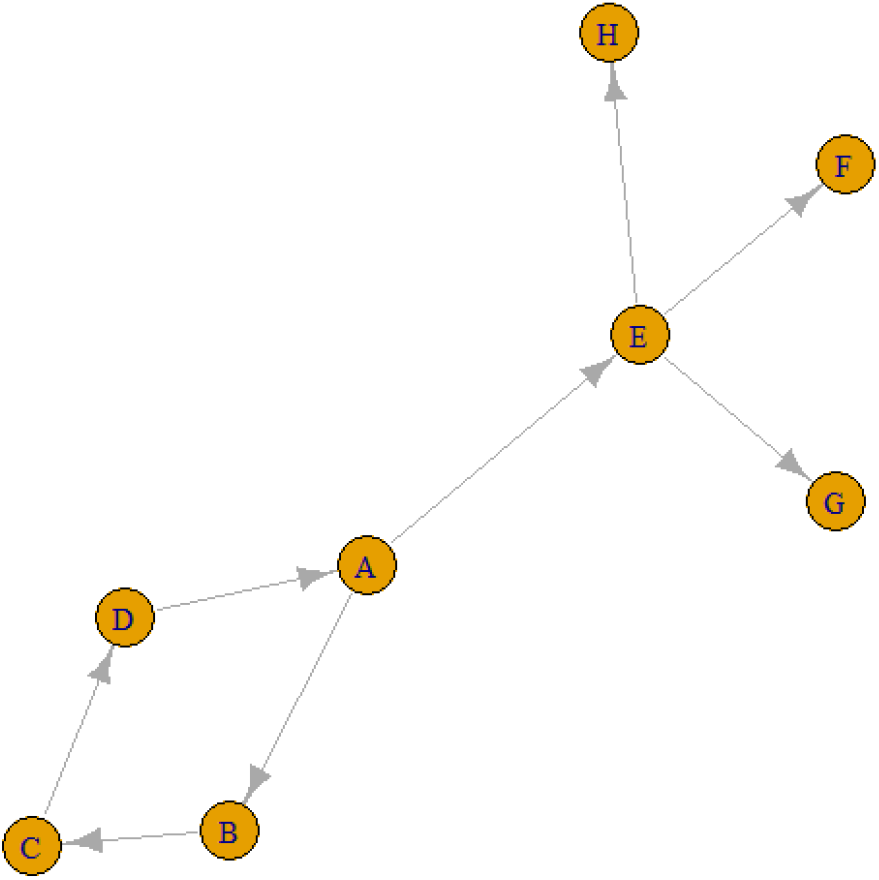
A transmission graph

It is also possible to find all the paths from any node to other nodes using these graphs. For example, according to the line above, there is a single ABC-shaped path leading to C in A. Similarly, from A to F can also be reached via the AEF pathway, and the number of jumps in both the ABC pathway and the AEF pathway is two.

> library(igraph)

> el <- cbind(c(“A”,”B”,”C”,”D”,”E”,”E”,”E”,”A”),

+ c(“B”,”C”, “D”, “A”,”F”,”G”,”H”,”E”))

> g <- graph.edgelist(el, directed=TRUE)

> all_simple_paths(g,”A”,c(“C”,”F”))

[[1]]

+ 3/8 vertices, named, from 307e42f:

[1] A B C [[2]]

+ 3/8 vertices, named, from 307e42f:

[1] A E F

Different personal contact network structures can reduce R, which represents how many individuals are infected by each carrier. “By introducing a social network approach, we propose that a decrease in R can simultaneously be achieved by managing the network structure of interpersonal contact” [31]. If we move from this point, we can think about different network structures that will reduce infectiousness in epidemics, considering that we may encounter different outbreaks in the future.

As it can be seen in Figure 4, “From a social network perspective, the shape of the infection curve is closely related to the concept of network distance (or path lengths), which indicates the number of network steps needed to connect two nodes” [31,32]

**Figure 4.**
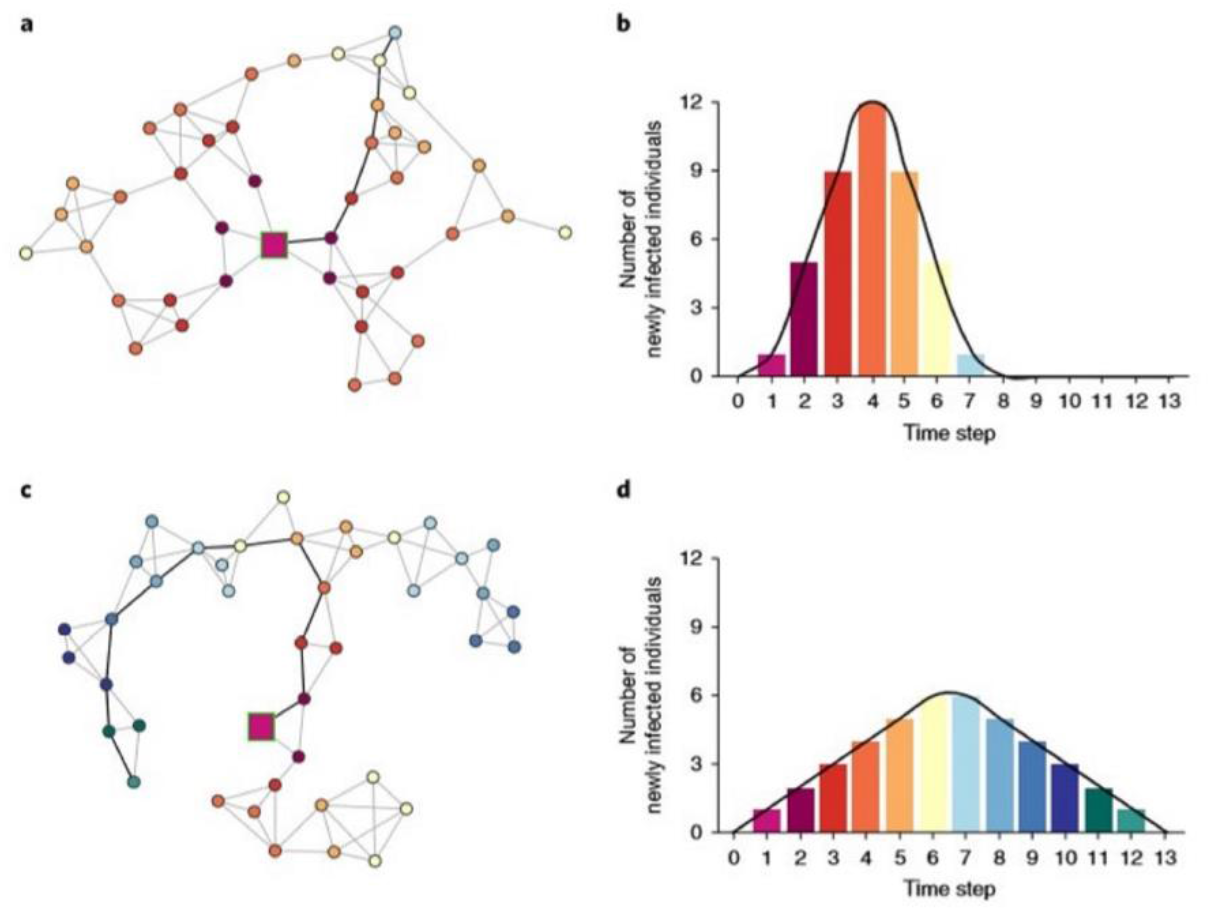
Two example networks. a-c with the same number of nodes and ties [31]

In Figure 4, two example networks. a-c, have the same number of nodes (individuals) and ties (social interactions) but different structures (shorter path lengths in a and longer path lengths in c), which imply different infection curves (b and d, respectively). Bold ties highlight the shortest infection path from the infection source to the last infected individual in the respective networks. Network node colour indicates at which step a node is infected and maps onto the colours of the histogram bars [31]

In order to obtain such a network, first we need to have a probability distribution regarding the number of people a person can infect the disease. When we have such a transmission probability distribution about the probabilities that a person can transmit the disease to how many people, we can draw a social network-like COVID-19 transmission graph by generating random variables about how many people can transmit the disease at each stage.

> library(igraph)

> g=graph(edges=c(1,2,1,3,1,4,2,5,2,6,2,7,2,8,

+ 3,9,3,10,3,11,4,12,12,13,12,14,12,15,12,16,10,17,10,18,

+ 9,19,9,20,9,21,5,22,6,23,6,24,6,25,7,26,8,27,8,28,

+ 11,29,11,30,11,31,11,32,11,33,11,34), n=34, directed=T)

> plot(g)

Figure 5 and 6 display us a COVID-19 transmission graph using simulated discrete Pareto distribution values. And in Figure 6 we can see the first and second stages of this graphs. In the third stage shape of the graph transforms into Figure 5.

**Figure 5.**
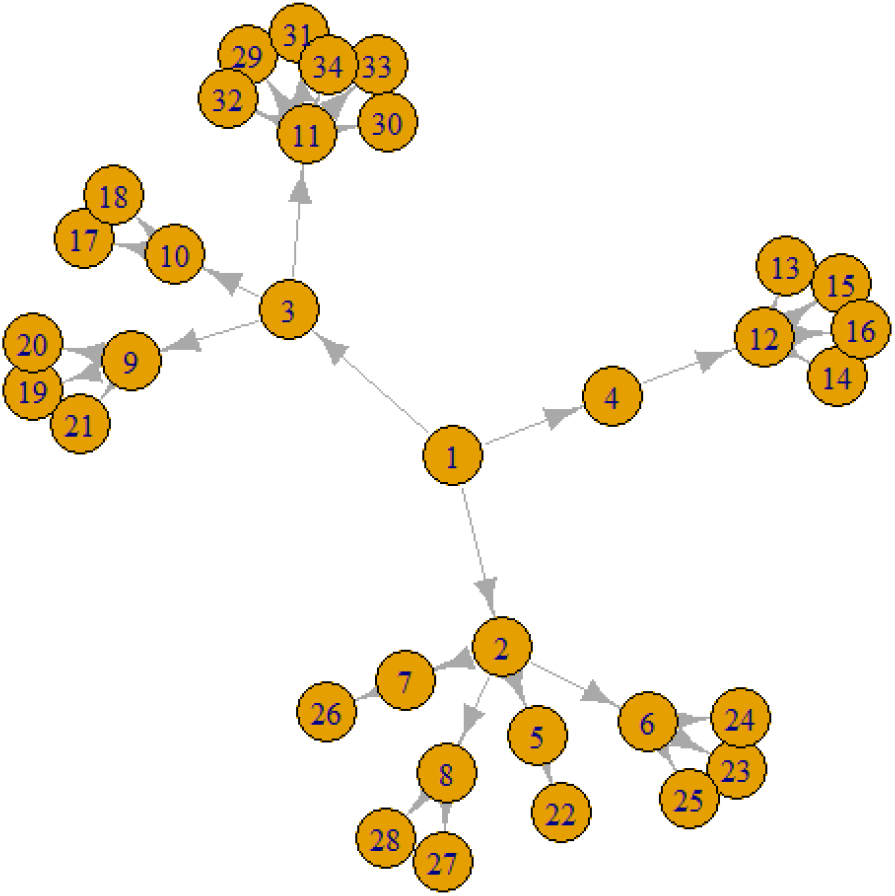
COVID-19 transmission graph using simulated discrete Pareto distribution values

**Figure 6.**
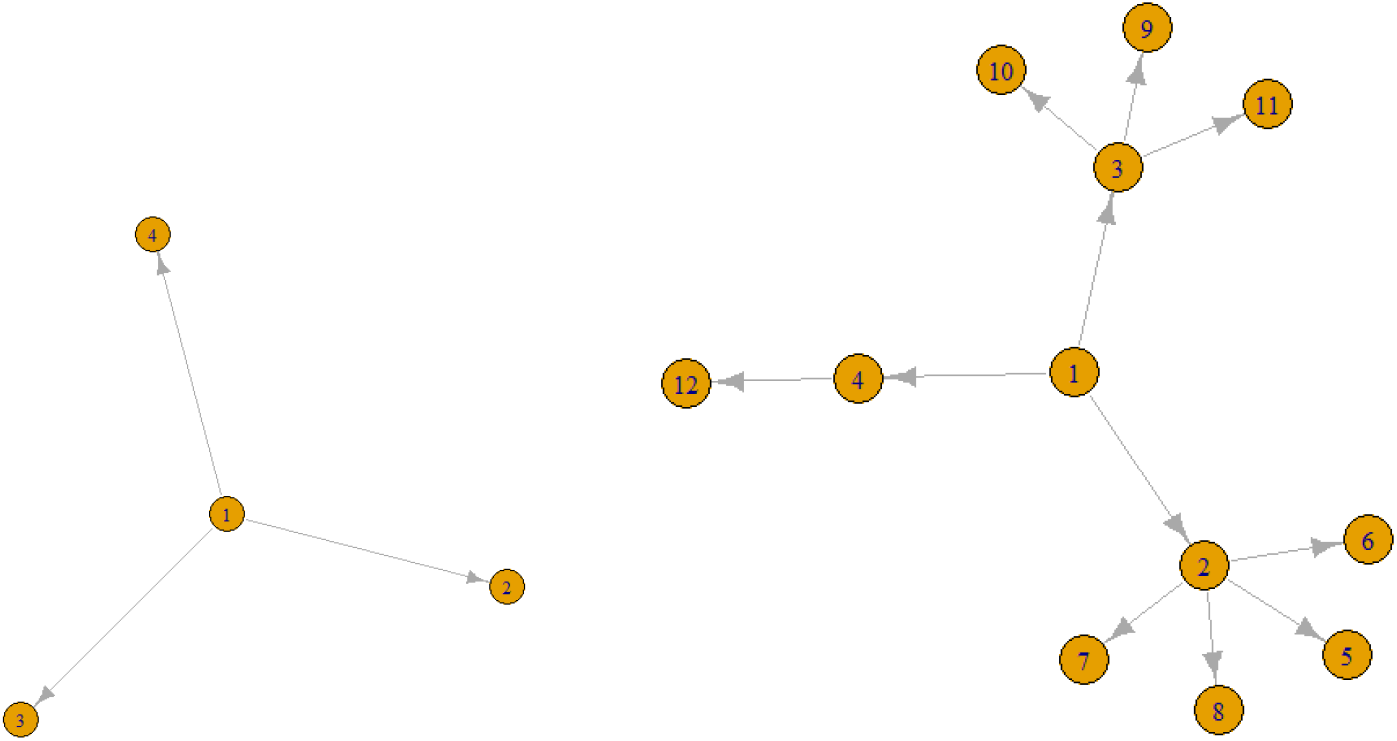
The first (on the left) and second stages (on the right) of COVID-19 transmission graphs

In figure 7, network structure of COVID-19 infections in India until March 13, 2020, reveals that most transmission has occurred from individuals with recent travel history abroad (node 0) [33].

**Figure 7:**
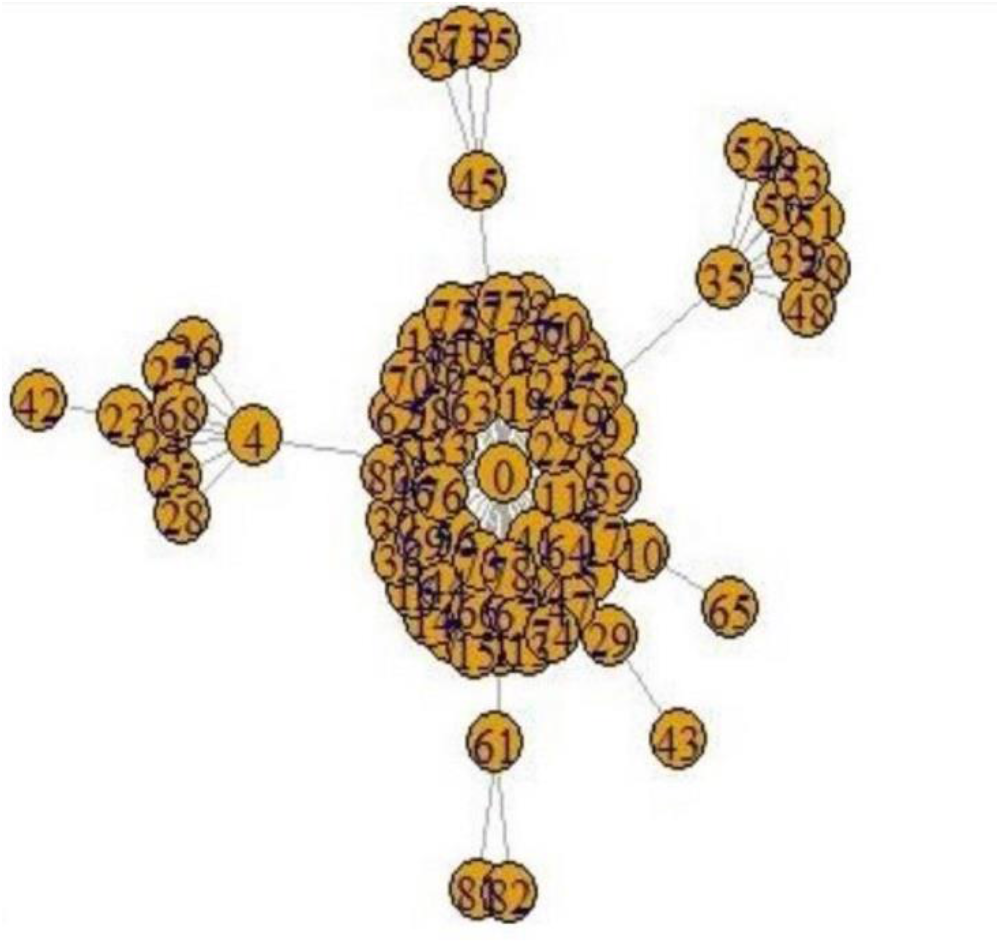
Network structure of COVID-19 infections in India until March 13, 2020 [33]

It is really interesting that similarities between our Figure 5 and Figure 7 of COVID-19 infections in India until March 13. Besides, Figure 7 of India has been drawn using real data. We know that, “Typically the network structure is inferred from indirect, incomplete, and often biased observations. Specification of an adjacency matrix is even more difficult when the underlying network is dynamic” and another centrality measure named expected force has been offered for additional advantages over existing spreading power and centrality measures [34]. But in our study we will use eigenvalue centrality measure to determine which nodes are super-spreaders. And as a result we have found that Node 3 is a super-spreader, as seen in figure 5.

> eigen_centrality(g)

$vector

[1] 0.52856114 0.47925178 **0.80872093** 0.23798419 0.18863564 0.25935484

[7] 0.18863564 0.21841342 0.43765240 0.36856516 1.00000000 0.15849962

[13] 0.05490112 0.05490112 0.05490112 0.05490112 0.12766364 0.12766364

[19] 0.15159409 0.15159409 0.15159409 0.06533964 0.08983536 0.08983536

[25] 0.08983536 0.06533964 0.07565407 0.07565407 0.34638012 0.34638012

[31] 0.34638012 0.34638012 0.34638012 0.34638012

## DISCUSSION

This study, which is conducted within the framework of interdisciplinary approach, focuses mainly on two purposes. In the first of these purposes transforming a transmission distribution of statistics and epidemiology into a transmission network of network science is aimed; In the second one, we try to determine who the super-spreaders are by using this network.

It is not appropriate to express the transmission distribution of this disease with an average R_0_ because this distribution is a power law descending from left to right. But at the beginning of the COVID-19 outbreak, this mistake has been made in the analysis. Besides, we know that different methods for R_0_ give different results and most of the times in scientific articles which method has been used is not mentioned.

And the second mistake is not to consider the importance of super-spreaders in this distribution. Looking at the outbreaks in history, it can be seen that the phenomenon of super-spreaders is not new and spanning about 2,500 years and the distribution of the victims of infectious diseases is extremely fat-tailed. Most of the times, the fact that 80% of the infection is carried out by a 20% group is often not considered and most of the analysis begin with a R_0_ reproduction number. In fact, we should add that there is a connection between these two errors and that this is a single error. The super-spreaders are the nodes in a network that can maximize their impacts on other nodes, in the case of virus transmission n. Although it is still unclear why certain individuals infect disproportionately large numbers of secondary contacts, the fact that 80% of the infection is carried out by a 20% group is important. But maybe it is necessary to add that some research tells us that 80% of secondary transmissions may have been caused by a small fraction of infectious individuals (~10%).

COVID-19 transmission is strongly heterogeneous, while a high degree of individual-level variation in the transmission of COVID-19 has been expressed and consensus range of R_0_ was found within the interval of 2-3. On the other hand, in Wuhan, R_0_ median daily reproduction number changes from 2.35 to 1.05 in only one week. From the results of another research we learn that R_0_ is between 2.6-3.2 for Republic of Korea and 2.6-3.3 for Italy and also these results falsify the above mentioned so called consensus.

We have to underline that point, determination of transmission probability distribution is a very important point in the analysis of the epidemic and determining the precautions to be taken. We know that in such a case network structure is inferred from indirect, incomplete, and often biased observations. As the main difficulty in establishing such a link is the lack of data and information due to the very recent COVID-19 that’s why required data are generated by simulation. After transforming a transmission distribution into a transmission network and having such a graph, also we may compute many network measures and use this measures in our decision process.

Disease transmission networks have the motifs of transmission stories. One of the most important ways to avoid contamination is to have information about how this transmission happens. From a social network perspective, the shape of the infection curve is closely related to the concept of network distance (or path lengths), which indicates the number of network steps needed to connect two nodes.

Contact networks and disease transmission networks are different from each other. Incoming and outcoming edges can be any number in a contact network but in a transmission network the number of incoming edge always must be one and outcoming edge can be any number like contact networks.

In our study, we have used eigenvalue centrality measure to determine which nodes are super-spreaders. In fact, the issue does not end at this point and is just beginning, because only if the biological, social and genetic features of the determined super-spreaders can be determined, only then can these people be prevented from accelerating the outbreak.

## Data Availability

Data derivation process and codes are accessible in the manuscript,

